# Safety and potential benefits of acute intermittent hypoxia in people with chronic traumatic brain injury

**DOI:** 10.1101/2025.09.24.25336487

**Authors:** Ekaterina Delikishkina, Alexander J. Barry, Julio C. Hernandez-Pavon, Julia Heilberg, Rhea Rahim, Sonia Singh, Guillermo Tafur, Leah O’Shea, Michael Guthrie, Elliot Roth, W. Zev Rymer, Jordan Grafman

**Affiliations:** Shirley Ryan AbilityLab, Chicago, IL, USA; Feinberg School of Medicine, Northwestern University, Chicago, IL, USA

**Keywords:** traumatic brain injury, TBI, acute intermittent hypoxia, AIH, neuroplasticity, recovery of function

## Abstract

**Background:** Acute intermittent hypoxia (AIH) was recently demonstrated to improve motor and cognitive function in several patient populations, including incomplete spinal cord injury, stroke, multiple sclerosis and mild cognitive impairment.

**Objective:** Our clinical trial aimed to establish if AIH can be safely administered in patients with traumatic brain injury (TBI) and to collect preliminary data about its potential efficacy for treating motor, cognitive and affective sequelae of TBI.

**Methods:** Twelve volunteers with chronic TBI underwent four AIH sessions conducted on separate days, in which they were exposed to fifteen 30-60-s hypoxic episodes interspersed with 60-90 s of breathing ambient air. Inspired oxygen (O_2_) concentration during hypoxic episodes was gradually reduced from 21% (equal to ambient air, sham), to 17%, 13%, and 9%, over the course of four sessions. Neuropsychological and motor tests were administered on days before and after AIH, as well as 60 min after each AIH session. In addition, transcranial magnetic stimulation (TMS) was applied to the hand motor area 45 min after the first (21% O_2_) and last (9% O_2_) AIH session.

**Results:** All participants tolerated the AIH sessions well and were able to complete the entire protocol. No significant improvement in cognition or mood was noted after the AIH intervention. Motor performance gradually improved over the course of the study, but no significant changes in response to TMS were found in corticospinal excitability.

**Conclusions:** AIH dosage as low as 9% O_2_ appears safe to use in chronic TBI, but its potential benefits remain to be investigated.

## Introduction

Traumatic brain injury (TBI) is a leading cause of long-term disability worldwide. Of the 20 million people who experience a TBI every year globally (Yan et al., 2025), many never achieve full recovery. Notably, long-lasting functional impairments are reported not only in individuals with moderate-to-severe injuries, but also in those who suffered a mild TBI or a concussion (Nelson et al., 2023). These impairments present a challenge for patients returning to work or school and impose difficulties on their everyday personal and social lives. While many rehabilitation therapies are directed at restoring sensory, motor and cognitive functions of affected individuals, none are designed to directly modulate neuroplasticity processes following injury.

A novel therapeutic tool called acute intermittent hypoxia (AIH), which consists in repeated reduction of oxygen in inspired air, has been recently introduced. AIH was shown to promote the synthesis of brain-derived neurotrophic factor (BDNF) that facilitates neuroplasticity (Baker-Herman et al., 2004; Satriotomo et al., 2016), and has been utilized to improve motor function in several patient populations, including those with incomplete spinal cord injury (SCI; Afsharipour et al., 2023; Alashram, 2025; Christiansen et al., 2021; Pearcey et al., 2024; Sandhu et al., 2021), stroke (Pearcey et al., 2025), and multiple sclerosis (Sandhu et al., 2024). According to several recent reports, AIH may also aid the cognitive rehabilitation of individuals with multiple sclerosis (Sandhu et al., 2024) and mild cognitive impairment (Serebrovska et al., 2019; Wang et al., 2020), as well as potentially serve as a tool for improving overall cognitive performance in older adults (Bayer et al., 2017; Schega et al., 2013, 2016).

We set out to study the effects of AIH on people with TBI. The three main aims of the study consisted in (1) determining the safety of administering AIH in subjects with TBI, (2) collecting preliminary data about the potential efficacy of this therapeutic tool for improving motor, cognitive and affective functioning of TBI patients, and (3) assessing the effect of AIH on corticospinal excitability.

## Methods

### Participants

Twelve individuals with a history of a single mild to moderate chronic TBI (8 female; 44.6±12.3 years old, range: 28–63 years; 41.4±27.4 months since injury, range: 11–115 months) were recruited to participate in the study. The participants’ average score on the Ohio State University TBI Identification Method Interview Form (Corrigan & Bogner, 2007), the TBI severity assessment scale, was 2.67±0.65 (range: 2–4). On this scale, a score of 2 corresponds to “mild TBI with no loss of consciousness (LOC)”, 3 corresponds to “mild TBI with LOC up to 30 min”, and 4 corresponds to “moderate TBI with LOC between 30 min and 24 h”. All participants reported English as their primary language and completed at least a high school education (17.3±2.5 years, range: 12–20). All were self-reported right-handers, with an average laterality quotient score equal to 90.6±17.0 (range: 50–100), as assessed by the Edinburgh Handedness Inventory Short Form (Veale, 2014). Demographic and clinical information for individual participants is reported in Table S1.

### Procedure

The study protocol was approved by the Northwestern University Institutional Review Board (STU00213969) and registered on ClinicalTrials.gov (NCT04890639). All participants provided written informed consent.

All participants completed six research visits to the Shirley Ryan AbilityLab between 2022 and 2024. The visits included four AIH treatment sessions (Visits 2-5), as well as the pre- and post-treatment assessments of motor, cognitive and affective function (Visits 1 and 6 respectively). All visits were scheduled at least 24 hours apart, with an average interval of 2.55±0.33 days (range: 1–6 days). One patient interrupted her participation after two visits for personal reasons but was reenrolled 25.3 weeks later at her request and completed the entire protocol starting from the baseline assessment.

### Medical screening

Prior to starting the AIH treatment, all recruited participants underwent a medical screening, which included a resting 12-lead EKG and brain MRI (3T Siemens MAGNETOM Prisma scanner with a 64-channel head coil, Erlangen, Germany), followed by a physical exam to confirm they had adequate cardiac and pulmonary fitness to participate in the study. Additionally, women of childbearing potential had blood drawn to confirm negative pregnancy. Brain MRI scan was repeated in Visit 6.

### AIH administration

During each of the four AIH interventions administered in Visits 2 through 5, the patients received brief bouts of reduced oxygen (O_2_), that they inhaled through a non-rebreathing face mask. The gas mixture was delivered using a customized Hyp-123 device (Hypoxico, Gardiner, NY), and O_2_ concentrations were monitored using a Maxtec Handi+ oxygen analyzer (Maxtec, Salt Lake City, UT). Participants’ peripheral oxygen saturation levels (SpO_2_) and heart rate (HR) were monitored in real time with a Nonin 7500 pulse oximeter (Nonin Medical Inc., Plymouth, MN). SpO_2_ and HR readings were sampled at a 100-Hz rate and logged in an Excel spreadsheet using a custom software written in C#. Blood pressure was measured at baseline, during and immediately after the AIH session. O_2_ concentration in the inhaled gas mixture was gradually reduced over the course of four AIH treatment sessions, starting from 21% O_2_ (sham session with room air supplied during hypoxia episodes, target SpO_2_ = 95%) in Visit 2, to 17% O_2_ (target SpO_2_ = 92%) in Visit 3, 13% O_2_ (target SpO_2_ = 87%) in Visit 4, and 9% O_2_ (target SpO_2_ = 82%) in Visit 5. Each AIH session consisted of 15 two-minute cycles, which included a 30-60-s hypoxia episode (with the duration of each episode adjusted depending on how long it took to bring the participant’s SpO_2_ down to the target value for a given session), followed by 60-90 s of normoxia, during which we ensured participants’ SpO_2_ recovered to their baseline level.

### Behavioral outcome measures

The same behavioral testing protocol was administered in Visit 1 and Visit 6. The assessment battery included standardized neuropsychological tests evaluating various cognitive functions (Delis-Kaplan Executive Function System Verbal Fluency Test, Repeatable Battery for the Assessment of Neuropsychological Status Update, California Verbal Learning Test–II, Trail Making Test Parts A and B), tests of fine motor control and coordination (Finger Tapping Test, Grooved Pegboard Test), computerized tests of implicit motor learning (Serial Reaction Time Task) and reward-related motivation (Effort Expenditure for Rewards Task), as well as the questionnaire assessing depressive symptoms (Beck Depression Inventory–II). In addition, a brief assessment, consisting of the Finger Tapping Test, Grooved Pegboard Test, Rey Auditory Verbal Learning Test, and a single-item Visual Analog Mood Scale, was administered 60 min after the completion of each AIH treatment in Visits 2 through 5. A detailed description of each behavioral task is provided in Table S2.

### Transcranial magnetic stimulation (TMS)

To assess the potential changes in corticospinal excitability associated with the AIH intervention, transcranial magnetic stimulation (TMS) was used to investigate motor evoked potentials (MEPs). Motor mapping with neuronavigation (Localite, St Augustin, Germany) using individual high-resolution T1-weighted images of the brain (TR = 2 s, TE = 2.99 ms, FA = 8°, FOV = 256 × 256 mm, 208 slices, TI = 900 ms) was completed after MRI in Visit 1. TMS was applied using a MagPro X100 with a MagOption stimulator and a C-B60 figure-of-eight coil (MagVenture, Farum, Denmark). We first found the resting motor threshold (rMT) for the left primary motor cortex (M1) for the contralateral (dominant) first dorsal interosseous (FDI) muscle. The rMT was determined as the lowest TMS intensity evoking MEPs with an amplitude ≥ 50 µV peak-to-peak in at least five out of ten consecutive trials (Rossini et al., 1994). For MEP recordings, 20 biphasic TMS pulses were administered to the left M1 at 120% rMT while MEPs were recorded from the right FDI (Bittium, Kuopio, Finland). The interstimulus interval (ISI) between pulses was set to 5 s. The MEPs were recorded 45 min after completing the AIH intervention in Visit 2 (21% O2, sham) and Visit 5 (9% O2). MEPs were averaged across trials, and peak-to-peak amplitudes between 20 and 40 ms after TMS pulses were measured to assess the effects of AIH.

The procedures and assessments involved in the six study visits are summarized in Table 1.

**Table 1.**
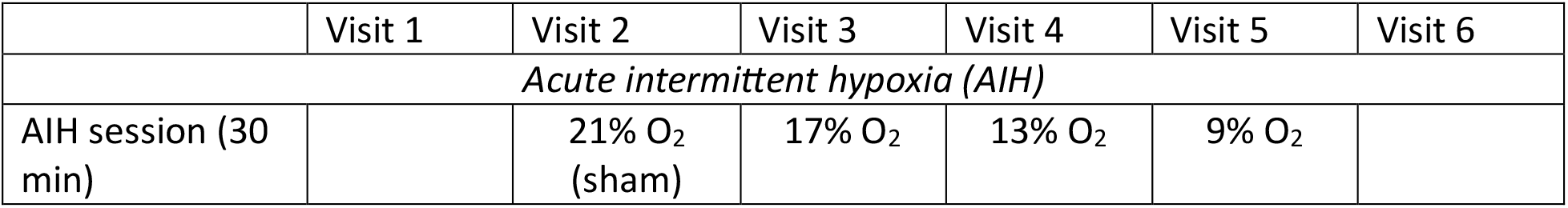
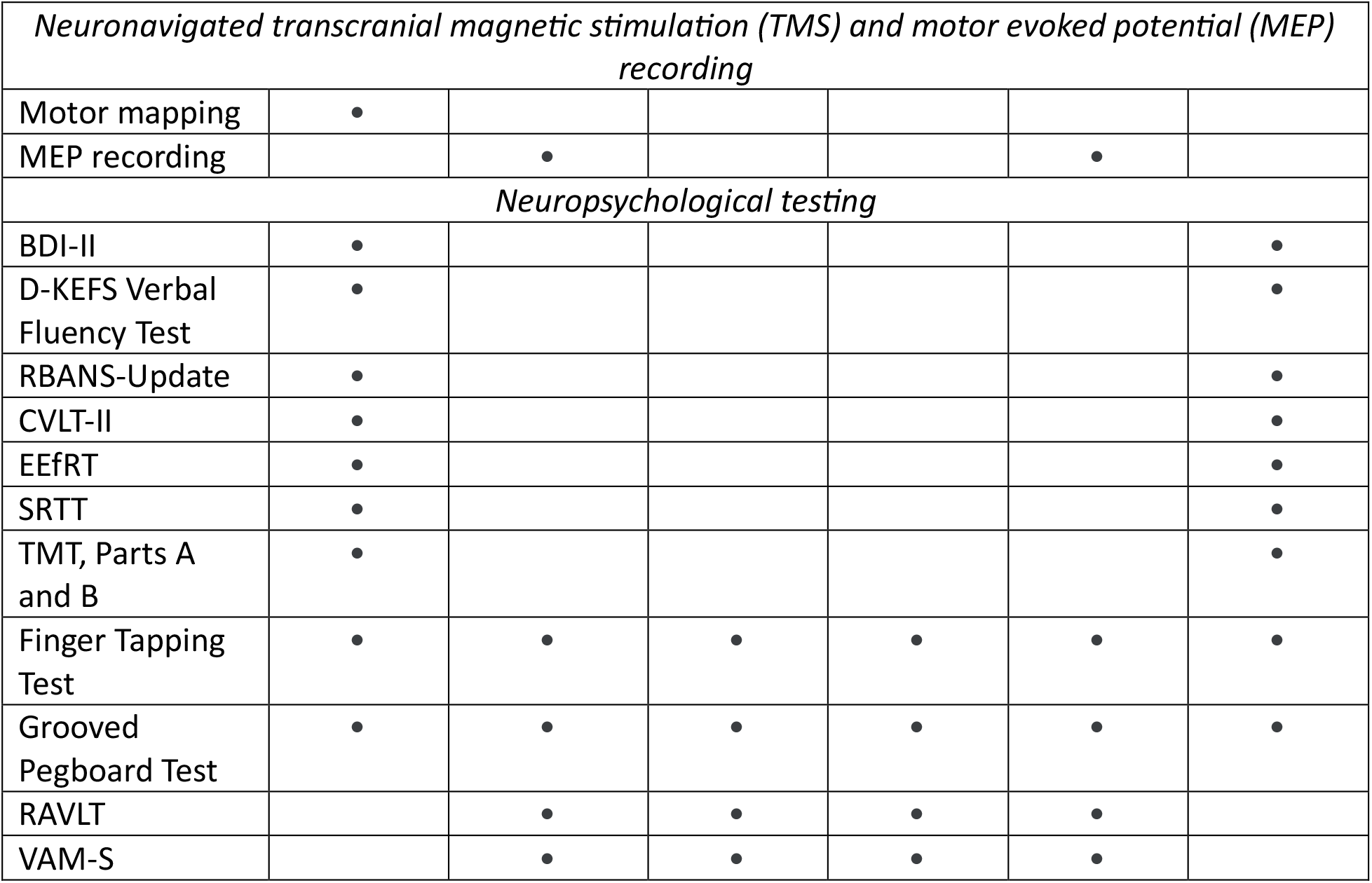
Experimental procedures and tests completed as part of the study. BDI-II = Beck Depression Inventory–II; D-KEFS = Delis-Kaplan Executive Function System; RBANS = Repeatable Battery for the Assessment of Neuropsychological Status Update; CVLT-II = California Verbal Learning Test–II; EEfRT = Effort Expenditure for Rewards Task; SRTT = Serial Reaction Time Task; TMT = Trail Making Test; RAVLT = Rey Auditory Verbal Learning Test; VAM-S = Visual Analog Mood Scale, single-item.

|  | Visit 1 | Visit 2 | Visit 3 | Visit 4 | Visit 5 | Visit 6 |
| --- | --- | --- | --- | --- | --- | --- |
| *Acute intermittent hypoxia (AIH)* |  |  |  |  |  |  |
| AIH session (30 min) |  | 21% O<sub>2</sub><br>(sham) | 17% O<sub>2</sub> | 13% O<sub>2</sub> | 9% O<sub>2</sub> |  |

| *Neuronavigated transcranial magnetic stimulation (TMS) and motor evoked potential (MEP) recording* |  |  |  |  |  |  |
| --- | --- | --- | --- | --- | --- | --- |
| Motor mapping | • |  |  |  |  |  |
| MEP recording |  | • |  |  | • |  |
| *Neuropsychological testing* |  |  |  |  |  |  |
| BDI-II | • |  |  |  |  | • |
| D-KEFS Verbal Fluency Test | • |  |  |  |  | • |
| RBANS-Update | • |  |  |  |  | • |
| CVLT-II | • |  |  |  |  | • |
| EEfRT | • |  |  |  |  | • |
| SRTT | • |  |  |  |  | • |
| TMT, Parts A and B | • |  |  |  |  | • |
| Finger Tapping Test | • | • | • | • | • | • |
| Grooved Pegboard Test | • | • | • | • | • | • |
| RAVLT |  | • | • | • | • |  |
| VAM-S |  | • | • | • | • |  |

#### Statistical analysis

Vital signs data (HR, SpO_2_) were preprocessed in MATLAB R2020a (MathWorks, Natick, MA). For each cycle in an AIH session, the highest HR and the lowest SpO_2_ readings were identified; then, the average highest HR (max HR) and lowest SpO_2_ (min SpO_2_) across all cycles were computed per session.

Across-session comparisons of behavioral test performance, vital signs (min HR, max SpO_2_) and MEPs were conducted using IBM SPSS Statistics v. 29.0, with the significance level set at α = .05. Normality of data was assessed using the Shapiro-Wilk test (α = .05). For normally distributed variables, means (*M*) and standard deviations (*SD*) were calculated, and groups were compared using a two-tailed paired-samples *t*-test or a repeated measures (RM) ANOVA. Mauchly’s test was used to check the RM ANOVA’s assumption of sphericity; when violated (*p* < .05), the Greenhouse-Geisser correction was applied. For non-normally distributed data, medians (*Mdn*) and interquartile ranges (*IQR*) were calculated, and group comparisons were performed using the Wilcoxon signed-rank test or the Friedman test. Effect sizes were calculated as follows: Cohen’s *d* for *t*-tests, *r* for Wilcoxon tests, partial eta squared (*η*_*p*_^*2*^) for RM ANOVA, and Kendall’s *W* for Friedman tests. Post hoc tests for RM ANOVA and Friedman tests were conducted using paired-samples *t*-tests and Wilcoxon signed-rank tests respectively, with multiple comparisons corrected using the false discovery rate (FDR) method (Benjamini & Hochberg, 1995), as implemented in the R function *p*.*adjust*. Data visualizations were created using the ggplot2 package (Wickham, 2016) in R v. 3.6.3 (R Core Team, 2020).

## Results

### Participant exclusions and missing data

Three participants were excluded from TMS: one could not complete the required brain MRI scan due to claustrophobia, and two were unable to undergo TMS due to scheduling issues. One participant was excluded from all assessments probing manual dexterity (Finger Tapping Test, Grooved Pegboard Test, Serial Reaction Time Task, Effort Expenditure for Rewards Task, Coding task of the Repeatable Battery for the Assessment of Neuropsychological Status Update) in Visit 6 due to a right arm fracture sustained three days prior. Another participant was excluded from the Grooved Pegboard Test in Visit 6 due to a bruised left thumb. Two participants did not complete the Effort Expenditure for Rewards Task in Visit 6: one failed to make a choice between the easy and hard versions of the task in all trials during Visit 1, and the other was unable to complete the hard version of the task within the allotted time during Visit 1. Additionally, Visit 1 data from the Serial Reaction Time Task were not recorded for two participants due to a technical error. Finally, data from the Rey Auditory Verbal Learning Test administered in Visits 2 through 5 were excluded from analysis due to significant ceiling effects – the same word list was used across all four sessions, leading to substantial long-term memory contamination. As a result, seven out of twelve participants were able to recall all words in Visit 3, and ten participants were able to recall all words in Visit 4, precluding reliable estimation of within-session learning trajectories. All remaining data were submitted to statistical analyses.

The full output of normality tests, assumption checks, inferential statistics, and descriptive measures is provided in Table S3. Detailed results of post hoc comparisons for omnibus tests are presented in Table S4. For clarity and brevity, only FDR-corrected *p*-values, effect sizes, and group averages illustrating the direction of effects are reported in the main text.

#### Adverse events and vital signs

All participants tolerated the AIH sessions well, and no serious task-related adverse events (AEs) were noted. The only related AE was transient lightheadedness, reported by two participants during AIH administration in Visit 4 (13% O2) and by one participant in Visit 5 (9% O2). Although self-reported symptom changes were not formally assessed, several participants informally reported perceived improvements over the course of the study, including reduced “brain fog” and enhanced physical or cognitive performance (e.g., “better memory”, “improved tennis game”).

Averaged across subjects, max HR gradually increased over the course of four AIH treatment sessions (Figure 1A), as min SpO_2_ gradually declined (Figure 1B). A main effect of hypoxia dosage on max HR was significant (RM ANOVA: *p* = .044, η_p_^2^ = .27); post hoc tests revealed that this effect was driven largely by a significant increase of max HR in Visit 5 compared to Visit 2 (*p* = .012, *d* = −1.04) and to Visit 4 (*M* = 76.75; *p* = .012, *d* = −1.14). A main effect of hypoxia dosage on min SpO_2_ was strongly significant (RM ANOVA: *p* < .001, η_p_^2^ = .90); all post hoc tests were significant (all *p*s ≤ .026), confirming that even mild exposure to hypoxic air caused a significant decrease in participants’ SpO_2_.

**Figure 1.**
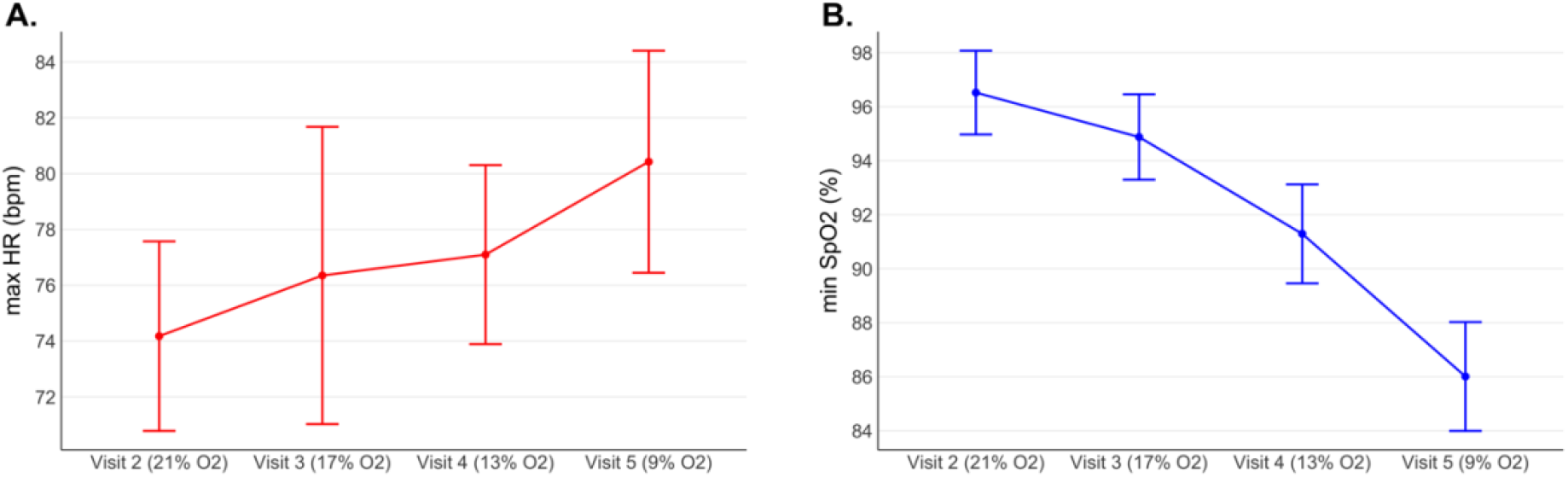
Maximum HR (**A**) and minimum SpO_2_ (**B**) averaged across subjects, over the four AIH treatment sessions. Error bars reflect 95% confidence intervals adjusted for the within-subject design using the Cousineau-Morey method (Cousineau, 2005; Morey, 2008).

#### Motor evoked potentials

Previous TMS studies in healthy volunteers (Christiansen et al., 2018) and patients with incomplete SCI (Christiansen et al., 2021) reported improved corticospinal excitability assessed with MEPs within 30-75 min following a single AIH session. To assess the potential AIH-induced motor plasticity in our TBI cohort, we compared MEP peak-to-peak amplitudes recorded from the right FDI 45 min after the sham AIH intervention in Visit 2 (21% O_2_) *vs* after completing the AIH protocol in Visit 5 (9% O_2_). Although average MEP amplitudes in our study were, contrary to expectations, lower in Visit 5 (*M* = 1180) compared to Visit 2 (*M* = 1625), this difference was not statistically significant (*p* = .312, *d* = .36; Figure S1). Moreover, the examination of individual MEP responses revealed marked across-subject variability, with some participants showing an increase and others showing a decrease in MEP amplitudes following AIH (Figure S2).

#### Behavioral tests

##### Motor function

As AIH was previously demonstrated to improve upper limb function in several patient populations, we hypothesized that the individuals with a chronic TBI enrolled in our study could show improved manual dexterity, as reflected by a gradual increase in the number of taps on the Finger Tapping Test and a decrease in the amount of time required to complete the Grooved Pegboard Test, proportional to the hypoxia dosage increase over the course of the study.

As demonstrated in Figure 2, the number of taps completed on the Finger Tapping Test, averaged across subjects, indeed gradually increased throughout the course of the study. A main effect of session on the number of taps was significant both for the right (Friedman test: *p* < .001, *W* = .45) and the left (RM ANOVA: p = .002, η_p_^2^ = .30) hands. However, only a few post hoc tests for the right hand remained significant after the FDR correction, including Visit 1 *vs* Visit 4 (*p* = .019, *r* = −.89), Visit 1 *vs* Visit 5 (*p* = .019, *r* = −.88), Visit 1 *vs* Visit 6 (*p* = .019, *r* = −.86), Visit 3 *vs* Visit 5 (*p* = .019, *r* = −.85) and Visit 3 *vs* Visit 6 (*p* = .027, *r* = −.79).

**Figure 2.**
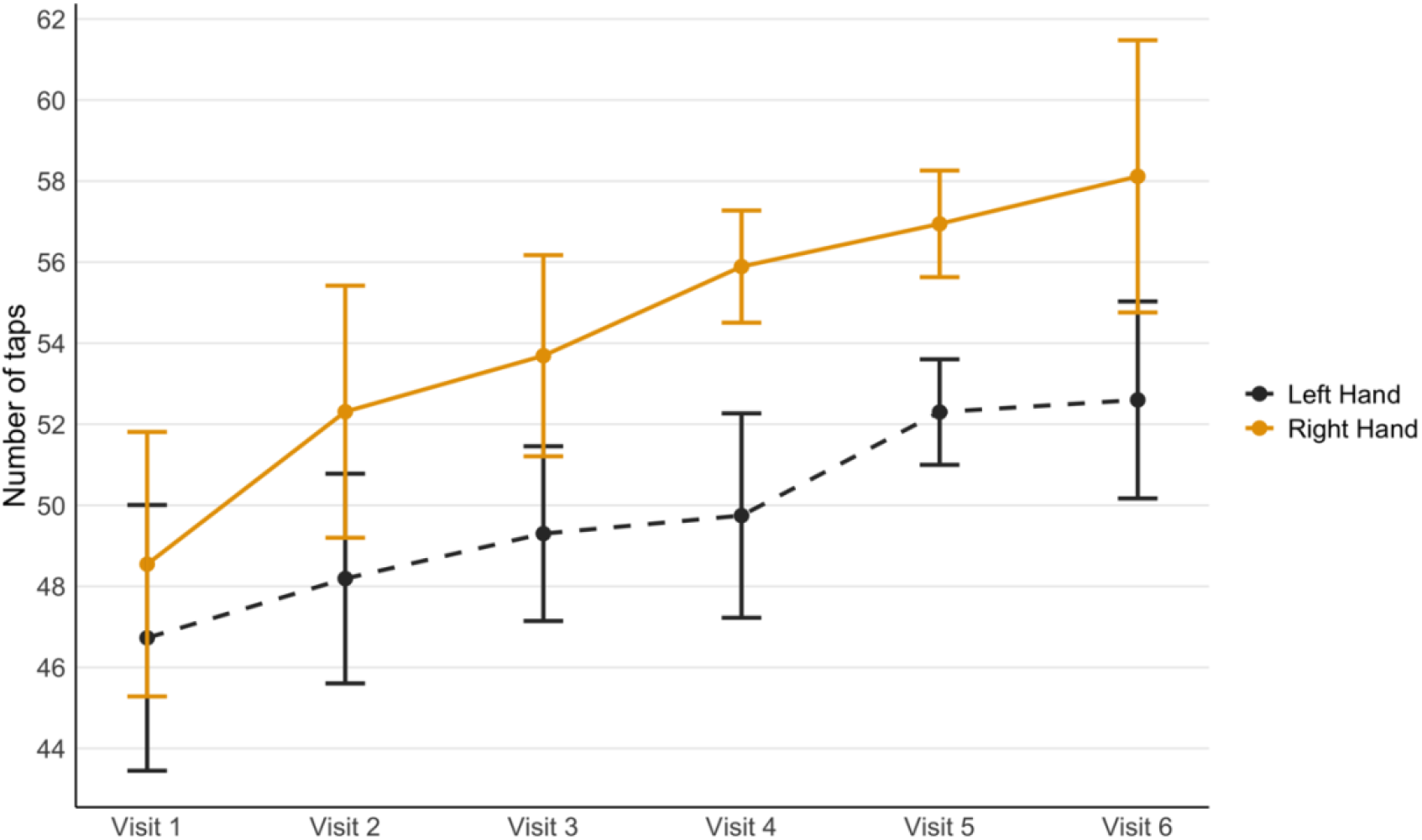
Finger Tapping Test scores (mean number of taps made with each hand across three experimental trials in a session), averaged across participants, over the course of six testing sessions. Error bars reflect 95% confidence intervals adjusted for the within-subject design using the Cousineau-Morey method (Cousineau, 2005; Morey, 2008).

The average time required to insert all pegs into the Grooved Pegboard decreased over the course of the study (Figure 3), and a main effect of session on test completion time was significant both for the right (Friedman test: *p* = .004, *W* = .34) and the left (RM ANOVA: *p* < .001, η_p_^2^ = .44) hands. Several post hoc tests survived the FDR correction, including Visit 1 *vs* Visit 2 (*p* = .034, *r* = .84), Visit 1 *vs* Visit 4 (*p* = .034, *r* = .82), Visit 1 *vs* Visit 5 (*p* = .034, *r* = .89) and Visit 1 *vs* Visit 6 (*p* = .034, *r* = .82) for the right hand, as well as Visit 1 *vs* Visit 2 (*p* = .026, *d* = 1.10), Visit 1 *vs* Visit 4 (*p* = .015, *d* = 1.31), Visit 1 *vs* Visit 5 (*p* = .015, *d* = 1.25) and Visit 1 *vs* Visit 6 (*p* = .015, *d* = 1.31) for the left hand.

**Figure 3.**
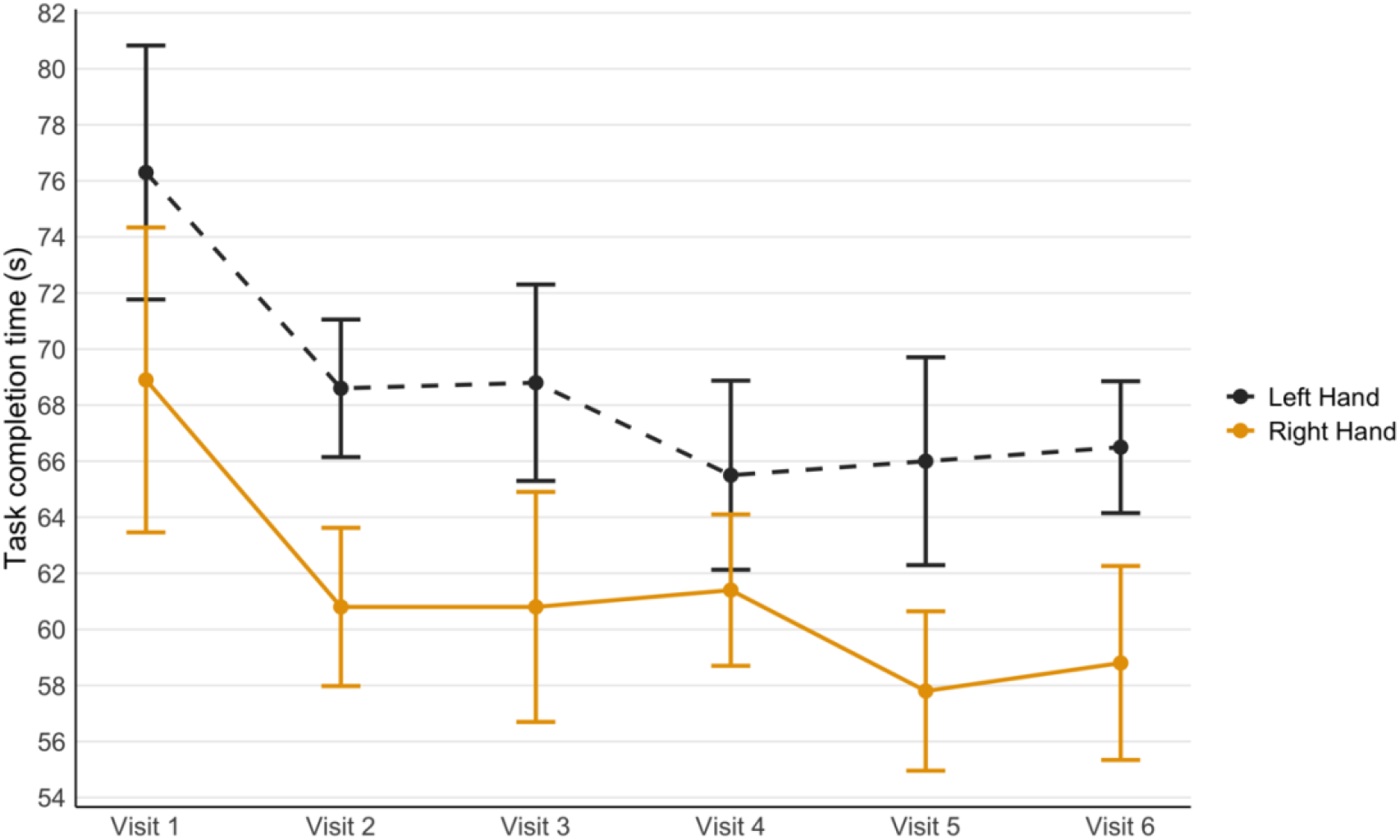
Performance on the Grooved Pegboard Test (total time in seconds required to complete the board with each hand), averaged across participants, over the course of six testing sessions. Error bars reflect 95% confidence intervals adjusted for the within-subject design using the Cousineau-Morey method (Cousineau, 2005; Morey, 2008).

Overall, the Finger Tapping and Grooved Pegboard Test results suggest that motor performance gradually improved over the course of the study. However, the fact that participants’ performance started improving before the first non-sham AIH intervention in Visit 3 suggests that this improvement must be at least partially attributed to task practice effects, rather than the effects of AIH treatment.

We also tested whether AIH affected participants’ implicit motor learning abilities by administering the computerized Serial Reaction Time Task in Visit 1 and Visit 6. In this task, reaction times (RTs) were recorded for cued key presses corresponding to an implicitly learned sequence, followed by a pseudorandom one. The underlying assumption was that successful acquisition of the learned sequence would lead to increased RTs during subsequent performance of the pseudorandom one, reflecting greater executive demands associated with inhibiting the previously learned pattern (Grafman et al., 1990). Yet, a comparison of the differences between average RTs in the pseudorandom and learned task blocks in Visit 1 (*Mdn* = .07) *vs* Visit 6 (*Mdn* = .05) revealed no significant change (*p* = .110, *r* = .53).

##### Cognitive function

To assess if AIH improved cognition in individuals with a chronic TBI, we compared participants’ performance on a variety of cognitive tests in Visit 1 and Visit 6.

Although the average number of correctly recalled items on most trials of the California Verbal Learning Test–II increased in Visit 6 compared to Visit 1, and the number of intrusions and repetitions declined (see Descriptive Statistics in Table S3), these changes were not statistically significant (all *p*s ≥ .053; see Inferential Statistics in Table S3). No significant differences were observed between Visit 1 and Visit 6 on any of the five cognitive domains or twelve individual tasks, nor on the total scores of the Repeatable Battery for the Assessment of the Neuropsychological Status Update (all *p*s ≥ .176). No significant differences in performance on either Trail Making Test Part A (*p* = .243, *r* = .35) or Part B (*p* = .157, *d* = .43) were found between Visit 1 and Visit 6.

The only cognitive test showing a significant change was Task 1 of the Verbal Fluency Test (Letter Fluency), with *lower* scores observed in Visit 6 (*M* = 46.7) compared to Visit 1 (*M* = 51.6; *p* = .003, *d* = 1.11). While this decline in performance following AIH was unexpected, it may have resulted from a confounding factor in the study design. Specifically, Verbal Fluency Task 1 was the first cognitive test administered in both sessions; however, whereas it followed TMS motor mapping in Visit 1, it was administered after a one-hour, no-task MRI scan in Visit 6. Several participants reported feeling drowsy immediately after the scan, which may have affected verbal output. Importantly, neither Verbal Fluency Task 2 (Category Fluency; *p* = 1, *d* = 0) nor Task 3 (Category Switching; *p* = .529, *d* = .19), nor any subsequent behavioral tasks showed similar declines, suggesting that this effect was task-specific and not generalizable to other cognitive or motor outcomes.

##### Affective function

To evaluate potential AIH-induced changes in reward motivation and effort-based decision-making, we administered the computerized Effort Expenditure for Rewards Task at Visit 1 and Visit 6. On each trial, participants chose between an easy and a hard version of the task based on the reward offered. The total proportion of hard-task choices, which was used as a measure of willingness to expend effort for rewards (Treadway et al., 2009), did not change significantly from Visit 1 (*M* = .35) to Visit 6 (*M* = .42; *p* = .295, *d* = −.37).

Comparison of participants’ scores on the Beck’s Depression Inventory–II revealed no significant change in depressive symptoms from Visit 1 (*Mdn* = 5.50) to Visit 6 (*Mdn* = 3.00; *p* = .057, *r* = .55).

A single-item Visual Analog Mood Scale administered after the AIH sessions in Visits 2 through 5 showed no significant effect of hypoxia dosage on mood (*p* = .268, η_p_^2^ = .11).

## Discussion

All 12 participants with TBI recruited into the study complied with the entire AIH treatment protocol without serious AEs at any treatment intensity level, suggesting that AIH involving 17%, 13% and 9% O_2_ hypoxia dosage is generally safe to use in people with mild to moderate TBI. Yet, although several individuals informally reported positive changes in their physical and cognitive abilities, including relief from brain fog and better memory, and no one reported feeling worse, subjective improvement in patients’ mood and cognition was not supported using standard statistical analyses. However, we caution that the absence of significant effects of treatment on the performance on tests evaluating cognitive and affective functioning could be explained by the small sample size or by insufficient intensity/duration of the treatment. It is also possible that the tests that were administered in Visit 6, more than 24 h after the last AIH treatment session, did not capture short-lived AIH-induced benefits. Importantly, confirming that hypoxia dosage as low as 9% appears safe for individuals with mild to moderate chronic TBI opens the potential for investigating its efficacy using more intensive protocols and exploring its use as a treatment for individuals in the acute phase of TBI recovery.

Notably, the participants gradually improved on the tests of motor function, including the Finger Tapping Test and Grooved Pegboard Test, as the treatment progressed, in line with the studies reporting improved upper limb function following AIH in patients with incomplete spinal cord injury (Afsharipour et al., 2023; Pearcey et al., 2024; Sandhu et al., 2021), stroke (Pearcey et al., 2025) and multiple sclerosis (Sandhu et al., 2024). However, no statistically significant changes were found in MEP amplitudes recorded from the FDI muscle of the dominant hand, suggesting that improvement on the motor tests could be attributed to task practice effects. Finally, it is possible that since participants were informed about the intensity of treatment in each session, this information could have contributed to the placebo effect. As a next step, conducting a large-scale double-blind study including a control group would help to avoid the potential confounds described above. The potential benefits of combining AIH with task-specific training targeting individual deficits (Vose et al., 2022; Welch et al., 2020), as well as cumulative effects of repeated AIH treatments, also remain to be explored.

## Data Availability

The data that support the findings of this study are available from the corresponding author upon reasonable request.

## Statements and Declarations

## Acknowledgements

We are deeply grateful to the study participants for generously contributing their time and effort to this research. We would like to thank Dr. Milap Sandhu for his advice on the study design and Isabel Jaffe for her assistance during AIH administration.

## Supplementary Materials

Supplementary figures and tables mentioned in this article are available online.

