## Supplementary figures and tables for "Safety and potential benefits of acute intermittent hypoxia in people with chronic traumatic brain injury"

### Supplementary Tables

**Table S1.** Demographic and clinical information about patients enrolled into the study. F = female, M = male; OSU = Ohio State University, TBI = Traumatic Brain Injury.

| Subject ID | Sex | Age range | Years of education | Handedness Laterality Quotient score | Months since injury | OSU TBI Identification Method score |
|---|---|---|---|---|---|---|
| SUB01 | F | 61-65 | 18 | 100 | 62 | 3 |
| SUB02 | M | 31-35 | 20 | 100 | 11 | 2 |
| SUB03 | F | 26-30 | 19 | 100 | 115 | 3 |
| SUB04 | F | 46-50 | 20 | 87.5 | 32 | 2 |
| SUB05 | F | 31-35 | 20 | 100 | 38 | 4 |
| SUB06 | F | 26-30 | 16 | 87.5 | 16 | 2 |
| SUB07 | F | 36-40 | 18 | 100 | 26 | 2 |
| SUB08 | M | 56-60 | 12 | 100 | 53 | 3 |
| SUB09 | M | 41-45 | 16 | 62.5 | 23 | 3 |
| SUB10 | F | 56-60 | 16 | 100 | 43 | 3 |
| SUB11 | M | 41-45 | 14 | 100 | 37 | 3 |
| SUB12 | F | 56-60 | 18 | 50 | 41 | 2 |

**Table S2.** Description of the behavioral tasks used to assess the potential neurocognitive changes associated with the AIH intervention.

| Behavioral task | Administered in visits | Brief description of the task | Dependent variable(s) | Sample size (N) |
|---|---|---|---|---|
| | | *Cognitive function* | | |
| Delis-Kaplan Executive Function System (D- | Visit 1<br>Visit 6 | In this standardized test, participants were given 60 seconds to name (1) as many words as they can beginning | total number of correct | 12 |

| | | | | |
|---|---|---|---|---|
| KEFS): Verbal Fluency Test (Delis et al., 2001) | | from a certain letter (three trials, e.g., letters F, A, and S), (2) as many words as they can belonging to a certain semantic category (two trials, e.g., Animals and Boys' Names), and (3) as many words as they can from two alternating semantic categories (e.g., Fruits and Furniture). Two different experimental lists were used (Standard and Alternate Form), with the order of presentation counterbalanced across participants. | responses (raw score) given on each of three tasks | |
| Repeatable Battery for the Assessment of Neuropsychological Status Update (RBANS-Update; Randolph, 2012) | Visit 1<br>Visit 6 | This standardized neuropsychological battery includes 12 tasks assessing five cognitive domains (Immediate Memory, Visuospatial/Constructional Abilities, Language, Attention, and Delayed Memory). Two different lists (Forms B and C) were used, with the order of presentation counterbalanced across participants. | raw scores on 12 tasks and five cognitive domains, as well as sum of index scores and total scaled score | 12 (11 on Coding Task) |
| California Verbal Learning Test–II (CVLT-II; Delis et al., 2000) | Visit 1<br>Visit 6 | The CVLT-II is a widely used standardized assessment of verbal learning and memory. A 16-item word list was read to participants for a total of five times (Trials 1-5), followed by an interference list (List B). Recall was assessed immediately after reading each list, as well as after a short and a long delay. Two different experimental lists were used (Standard and Alternate Form), with the order of presentation counterbalanced across participants. | number of correctly recalled items on each trial, including immediate free recall, short- and long-delay free and cued recall; number of intrusions and repetitions; number of recognition hits | 12 |

| | | | | |
|---|---|---|---|---|
| Rey Auditory Verbal Learning Test (RAVLT; Schmidt, 1996) | Visit 2<br>Visit 3<br>Visit 4<br>Visit 5 | The RAVLT is a standardized assessment of verbal learning and memory. A 15-item word list was read to participants for a total of five times (Trials I-V), followed by an interference list (Trial B), a short-delay (Trial VI) and a long-delay (Trial VII) free recall, as well as a long-delay forced-choice recognition (Trial VIII) of the target list. The same (and the only one available) experimental list was administered in all four sessions. | number of correctly recalled items on each trial, including immediate free recall, short- and long-delay free recall; number of intrusions and repetitions; number of recognition hits | 12 |
| Trail Making Test (TMT): Parts A and B (Reitan, 1958) | Visit 1<br>Visit 6 | This task probes attention, visual search and executive function. In both parts of the task, participants were asked to connect the circles randomly placed on a sheet of paper in sequential order as fast as they can, without lifting the pen. In Trail A, they needed to connect the numbers from 1 to 25 in ascending order (1-2-3, etc.). In Trail B, they needed to get from 1 to 13 by alternating numbers and letters (1-A-2-B, etc.). | time in seconds required to complete each Trail | 11 |
| | | *Reward and motivation* | | |
| Effort Expenditure for Rewards Task (EEfRT; Treadway et al., 2009) | Visit 1<br>Visit 6 | In this task, implemented in Psychtoolbox-3 (Brainard, 1997) for MATLAB (MathWorks, Natick, MA), participants were presented with a series of trials in which they were prompted to choose between an Easy and a Hard version of the task based on the probability they would receive a monetary reward in case of successful task completion (high – 88%, medium – 50%, low – 12%) and the amount of reward ($1.00 for completing the Easy task or an amount varying between $1.24 and $4.21 for completing | total proportion of Hard task choices made by participants | 9 |

| | | | | |
|---|---|---|---|---|
| | | the Hard task). Participants were asked to choose the task during a 5-s bid presentation at the beginning of a trial, during which they were shown the amount of potential reward and their probability of winning. If the participant did not make their choice within the allotted time, the computer assigned the task version for them. The Easy task consisted in pressing the "L" key on the keyboard with the right (dominant) index finger for 30 times within the 7-second time window. The Hard task consisted in pressing the "S" key with the left (non-dominant) pinky finger for 98 times within the 21-second time window. While performing the task, participants raced against the timer and saw a progress bar showing how close they were to completing the task. At the beginning of the experiment, participants did four practice trials to ensure they understood the task and were able to complete it within the allotted time. Trials in which participants did not make a task choice or took less than 200 ms to choose the task, as well as four practice trials, were excluded from the analysis. | | |
| *Mood* | | | | |
| Beck Depression Inventory–II (BDI-II; Beck et al., 1996) | Visit 1<br>Visit 6 | This is a widely used 21-item self-report questionnaire assessing the severity of depressive symptoms. Each question has four response options, scored from 0 to 3 (the higher the score, the more severe are depression symptoms). | total score on the questionnaire | 12 |
| Visual Analog Mood Scale (VAM-S; Folstein & Luria, 1973) | Visit 2<br>Visit 3<br>Visit 4<br>Visit 5 | In this simple test, participants were asked to draw a mark on a 10-cm line corresponding to their current mood (with 'very bad' on the left, 'very good' on the right and 'neutral' in the middle). | length of the segment between the leftmost part of the scale and | 12 |

| | | | | |
|---|---|---|---|---|
| | | | the mark made by the subject in cm | |
| *Motor function* | | | | |
| Finger Tapping Test (Reitan & Wolfson, 1985; Ruff & Parker, 1993) | Visit 1<br>Visit 2<br>Visit 3<br>Visit 4<br>Visit 5<br>Visit 6 | This test was used as a basic measure of motor performance and was administered with one hand at a time, starting with the dominant (right) one. Participants were asked to place their hand on a wooden board with a lever attached to it and to rest their index finger on the lever. Next, five ten-second trials were administered, in which participants had to press the lever as fast as they could, aiming to do as many taps as possible within the allotted time. The number of taps on each trial was recorded by the experimenter. The first two trials served as warm-ups and were excluded from analysis. If the difference between the best and the worst attempt in trials 3 through 5 exceeded 15 taps, the experiment was repeated for another five trials and only the data from the second attempt were used. | mean number of taps on trials 3 through 5, separately for each hand | 11 |
| Grooved Pegboard Test (Ruff & Parker, 1993) | Visit 1<br>Visit 2<br>Visit 3<br>Visit 4<br>Visit 5<br>Visit 6 | This test was used as a measure of manual dexterity and eye-hand coordination and consisted in inserting grooved pegs into a 5×5 board (Model 32025; Lafayette Instrument Company, 2015) as quickly as possible, without skipping any slots, starting from the top row. One hand was tested at a time, starting with the right (dominant) one. When using the right hand, the patients were asked to fill the board from left to right. When using the left hand, they had to go in the opposite direction, from right to left. | total time in seconds required to complete the task with each hand | 10 |
| Serial Reaction Time Task (SRTT; Nissen et al., | Visit 1<br>Visit 6 | This task was used as a measure of implicit motor learning. The MATLAB code for task presentation was adapted from Schorn & Knowlton (2021) and presented | difference in reaction times (RTs) between | 9 |

| | | | |
|---|---|---|---|
| 1987; Nissen & Bullemer, 1987) | | using Psychtoolbox-3. At the beginning of the experiment, participants were sat in front of a computer and were instructed to hover four fingers of their right (dominant) hand over four keyboard keys (“U”, “I”, “O”, “P”). At the beginning of the task, four horizontally aligned unfilled circles appeared on the blue screen. The circles would become filled with white color in sequence, one at a time. Participants were instructed to press the corresponding key (the leftmost circle corresponding to “U”, the second one to the left to “I”, etc.) as quickly and accurately as possible after receiving the cue. If they responded incorrectly or took over 1 s to respond, they heard a tone. In our version of the task, participants completed six blocks of the task, presented consecutively without a break. Each block consisted of 100 trials. In blocks 1 and 6 participants were presented with pseudorandom key sequences. In blocks 2 through 5 they received the same 10-item sequence, that was repeated for a total of ten times per block. Participants were not informed about the presence of a repeating pattern. Two different patterns were used in Visit 1 and Visit 6 to avoid the potential confounding by long-term memory recall. The inter-trial interval (ITI) was set to 500 ms. All trials were included in the analysis, regardless of accuracy. | block 6 (pseudorandom) and block 5 (repeated) — if participants successfully acquired the repeated pattern by block 5, its interruption in block 6 would cause an increase in RTs (Grafman et al., 1990) |

**Table S3.** The output of normality tests, assumption checks, inferential statistics, and descriptive measures for the tests administered in order to assess the potential physiological, motor, cognitive and affective changes associated with the AIH intervention.

| | Visit | Normality test (Shapiro-Wilk) | Descriptive statistics | Inferential statistics |
|---|---|---|---|---|
| **Vitals during AIH** | | | | |
| max HR | Visit 2 | *W*(12) = .917, *p* = .264 | *M* = 74.18, *SD* = 11.40 | Mauchly' test: $\chi^2(5)$ = 13.54, *p* = **.019**, Greenhouse-Geisser correction: ε = .542), RM ANOVA: *F*(1.63, 17.89) = 4.01, *p* = **.044**, $\eta_p^2$ = .27 |
| | Visit 3 | *W*(12) = .953, *p* = .675 | *M* = 76.26, *SD* = 11.62 | |
| | Visit 4 | *W*(12) = .928, *p* = .361 | *M* = 76.75, *SD* = 13.00 | |
| | Visit 5 | *W*(12) = .941, *p* = .513 | *M* = 80.43, *SD* = 13.79 | |
| min $SpO_2$ | Visit 2 | *W*(12) = .891, *p* = .120 | *M* = 96.52, *SD* = 1.93 | Mauchly's test: $\chi^2(5)$ = 9.27, *p* = .100, RM ANOVA: *F*(3, 33) = 95.45, *p* < **.001**, $\eta_p^2$ = .90 |
| | Visit 3 | *W*(12) = .989, *p* = 1 | *M* = 94.87, *SD* = 1.45 | |
| | Visit 4 | *W*(12) = .936, *p* = .452 | *M* = 91.09, *SD* = 2.69 | |
| | Visit 5 | *W*(12) = .915, *p* = .249 | *M* = 86.01, *SD* = 2.15 | |
| **Motor evoked potentials (MEP) from the right first dorsal interosseous (FDI) muscle following TMS** | | | | |
| MEP | Visit 2 | *W*(9) = .876, *p* = .142 | *M* = 1625.13, *SD* = 1185.61 | *t*(8) = 1.079, *p* = .312, *d* = .36 |
| | Visit 5 | *W*(9) = .956, *p* = .751 | *M* = 1179.86, *SD* = 825.37 | |
| **Behavioral testing** | | | | |
| **Finger Tapping Test** | | | | |
| Finger Tapping Test, right hand | Visit 1 | *W*(11) = .838, *p* = **.030** | *Mdn* = 49.30, *IQR* = 6.30 | Friedman test: $\chi^2(5)$ = 25.00, *p* < **.001**, *W* = .45 |
| | Visit 2 | *W*(11) = .912, *p* = .257 | *Mdn* = 52.70, *IQR* = 8.30 | |
| | Visit 3 | *W*(11) = .972, *p* = .907 | *Mdn* = 51.30, *IQR* = 15.00 | |
| | Visit 4 | *W*(11) = .912, *p* = .255 | *Mdn* = 54.30, *IQR* = 12.70 | |
| | Visit 5 | *W*(11) = .931, *p* = .426 | *Mdn* = 53.70, *IQR* = 14.70 | |
| | Visit 6 | *W*(11) = .878, *p* = .099 | *Mdn* = 55.70, *IQR* = 12.00 | |
| Finger Tapping Test, left hand | Visit 1 | *W*(11) = .953, *p* = .679 | *M* = 46.73, *SD* = 4.34 | Mauchly's test: $\chi^2(14)$ = 23.00, *p* = .070, *F*(5, 50) = 4.36, *p* = **.002**, $\eta_p^2$ = .30 |
| | Visit 2 | *W*(11) = .971, *p* = .894 | *M* = 48.19, *SD* = 8.39 | |
| | Visit 3 | *W*(11) = .977, *p* = .947 | *M* = 49.30, *SD* = 7.47 | |
| | Visit 4 | *W*(11) = .971, *p* = .896 | *M* = 49.75, *SD* = 9.35 | |

| | | | | |
|---|---|---|---|---|
| | Visit 5 | *W*(11) = .931, *p* = .418 | *M* = 52.30, *SD* = 7.63 | |
| | Visit 6 | *W*(11) = .969, *p* = .876 | *M* = 52.60, *SD* = 6.81 | |
| **Grooved Pegboard Test** | | | | |
| Grooved Pegboard Test, right hand | Visit 1 | *W*(10) = .807, *p* = **.018** | *Mdn* = 67.00, *IQR* = 11.00 | Friedman test: $\chi^2(5) = 17.14$, *p* = **.004**, *W* = .34 |
| | Visit 2 | *W*(10) = .928, *p* = .431 | *Mdn* = 61.00, *IQR* = 11.00 | |
| | Visit 3 | *W*(10) = .936, *p* = .512 | *Mdn* = 60.50, *IQR* = 16.00 | |
| | Visit 4 | *W*(10) = .962, *p* = .812 | *Mdn* = 62.00, *IQR* = 12.00 | |
| | Visit 5 | *W*(10) = .845, *p* = **.050** | *Mdn* = 56.00, *IQR* = 11.00 | |
| | Visit 6 | *W*(10) = .929, *p* = .435 | *Mdn* = 61.00, *IQR* = 14.00 | |
| Grooved Pegboard Test, left hand | Visit 1 | *W*(10) = .942, *p* = .581 | *M* = 76.30, *SD* = 14.08 | Mauchly's test: $\chi^2(14) = 10.46$, *p* = .746, RM ANOVA: *F*(5, 45) = 7.08, *p* < **.001**, $\eta_p^2 = .44$ |
| | Visit 2 | *W*(10) = .969, *p* = .883 | *M* = 68.60, *SD* = 8.81 | |
| | Visit 3 | *W*(10) = .948, *p* = .650 | *M* = 68.80, *SD* = 9.37 | |
| | Visit 4 | *W*(10) = .909, *p* = .274 | *M* = 65.50, *SD* = 8.09 | |
| | Visit 5 | *W*(10) = .983, *p* = .978 | *M* = 66.00, *SD* = 10.73 | |
| | Visit 6 | *W*(10) = .964, *p* = .830 | *M* = 66.50, *SD* = 9.61 | |
| **Serial Reaction Time Task (SRTT)** | | | | |
| Serial Reaction Time Task | Visit 1 | *W*(9) = .886, *p* = .182 | *Mdn* = .07, *IQR* = .17 | *Z* = 1.60, *p* = .110, *r* = .53 |
| | Visit 6 | *W*(9) = .772, *p* = .010 | *Mdn* = .05, *IQR* = .05 | |
| **California Verbal Learning Memory–II (CVLT-II)** | | | | |
| CVLT-II: Trial 1 Free Recall | Visit 1 | *W*(12) = .974, *p* = .945 | *M* = 5.25, *SD* = 2.09 | *t*(11) = −1.732, *p* = .111, *d* = −.50 |
| | Visit 6 | *W*(12) = .903, *p* = .173 | *M* = 6.25, *SD* = 2.45 | |
| CVLT-II: Trial 2 Free Recall | Visit 1 | *W*(12) = .915, *p* = .245 | *M* = 8.75, *SD* = 2.45 | *t*(11) = −.306, *p* = .766, *d* = −.09 |
| | Visit 6 | *W*(12) = .941, *p* = .517 | *M* = 9.00, *SD* = 2.99 | |
| CVLT-II: Trial 3 Free Recall | Visit 1 | *W*(12) = .926, *p* = .338 | *M* = 10.42, *SD* = 3.26 | *t*(11) = −1.217, *p* = .249, *d* = −.35 |
| | Visit 6 | *W*(12) = .952, *p* = .668 | *M* = 11.33, *SD* = 2.57 | |
| CVLT-II: Trial 4 Free Recall | Visit 1 | *W*(12) = .927, *p* = .354 | *M* = 11.33, *SD* = 2.81 | *t*(11) = −.442, *p* = .667, *d* = −.13 |
| | Visit 6 | *W*(12) = .953, *p* = .679 | *M* = 11.58, *SD* = 3.37 | |
| CVLT-II: Trial 5 Free Recall | Visit 1 | *W*(12) = .907, *p* = .198 | *M* = 12.50, *SD* = 2.81 | *t*(11) = −1.032, *p* = .324, *d* = −.30 |
| | Visit 6 | *W*(12) = .881, *p* = .091 | *M* = 13.00, *SD* = 2.63 | |
| | Visit 1 | *W*(12) = .964, *p* = .837 | *M* = 48.25, *SD* = 11.64 | |

| | | | | |
|---|---|---|---|---|
| CVLT-II: Trials 1-5 Free Recall | Visit 6 | *W*(12) = .933, *p* = .413 | *M* = 50.33, *SD* = 11.43 | *t*(11) = −.913, *p* = .381, *d* = −.26 |
| CVLT-II: List B Free Recall | Visit 1 | *W*(12) = .938, *p* = .468 | *Mdn* = 5.00, *IQR* = 3.00 | *Z* = .207, *p* = .836, *r* = .06 |
| | Visit 6 | *W*(12) = .849, *p* = **.036** | *Mdn* = 5.50, *IQR* = 3.00 | |
| CVLT-II: Short Delay Free Recall | Visit 1 | *W*(12) = .866, *p* = .058 | *M* = 10.75, *SD* = 4.03 | *t*(11) = −1.059, *p* = .312, *d* = −.31 |
| | Visit 6 | *W*(12) = .941, *p* = .507 | *M* = 11.50, *SD* = 3.43 | |
| CVLT-II: Short Delay Cued Recall | Visit 1 | *W*(12) = .909, *p* = .209 | *M* = 12.00, *SD* = 2.95 | *t*(11) = −.252, *p* = .806, *d* = −.07 |
| | Visit 6 | *W*(12) = .914, *p* = .233 | *M* = 12.17, *SD* = 3.54 | |
| CVLT-II: Long Delay Free Recall | Visit 1 | *W*(12) = .920, *p* = .284 | *M* = 11.00, *SD* = 3.77 | *t*(11) = 0, *p* = 1, *d* = 0 |
| | Visit 6 | *W*(12) = .918, *p* = .269 | *M* = 11.00, *SD* = 4.16 | |
| CVLT-II: Long Delay Cued Recall | Visit 1 | *W*(12) = .916, *p* = .255 | *M* = 11.75, *SD* = 3.31 | *t*(11) = −.143, *p* = .889, *d* = −.04 |
| | Visit 6 | *W*(12) = .884, *p* = .097 | *M* = 11.83, *SD* = 3.90 | |
| CVLT-II: Free Recall Intrusions, All Types | Visit 1 | *W*(12) = .899, *p* = .152 | *M* = 4.25, *SD* = 2.90 | *t*(11) = 2.169, *p* = .053, *d* = .63 |
| | Visit 6 | *W*(12) = .905, *p* = .185 | *M* =2.08, *SD* = 1.93 | |
| CVLT-II: Cued Recall Intrusions, All Types | Visit 1 | *W*(12) = .891, *p* = .123 | *Mdn* = 2.00, *IQR* = 4.00 | *Z* = 1.675, *p* = .094, *r* = .48 |
| | Visit 6 | *W*(12) = .576, *p* < **.001** | *Mdn* = 0.00, *IQR* = 2.00 | |
| CVLT-II: Total Intrusions, All Recall Trials, All Types | Visit 1 | *W*(12) = .928, *p* = .356 | *Mdn* = 5.00, *IQR* = 9.00 | *Z* = 1.730, *p* = .084, *r* = .50 |
| | Visit 6 | *W*(12) = .694, *p* < **.001** | *Mdn* = 2.50, *IQR* = 3.00 | |
| CVLT-II: Total Repetitions, All Recall Trials | Visit 1 | *W*(12) = .831, *p* = **.019** | *Mdn* = 2.00, *IQR* = 4.00 | *Z* = 1.279, *p* = .201, *r* = .37 |
| | Visit 6 | *W*(12) = .827, *p* = **.021** | *Mdn* = 1.00, *IQR* = 2.00 | |
| CVLT-II: Long Delay, Yes/No Recognition Hits | Visit 1 | *W*(12) = .792, *p* = **.045** | *Mdn* = 15.00, *IQR* = 3.00 | *Z* = .957, *p* = .339, *r* = .28 |
| | Visit 6 | *W*(12) = .857, *p* = **.008** | *Mdn* = 15.00, *IQR* = 4.00 | |
| **Delis-Kaplan Executive Function System (D-KEFS): Verbal Fluency Test** | | | | |
| D-KEFS Verbal Fluency: Task 1 (Letter Fluency) | Visit 1 | *W*(12) = .962, *p* = .813 | *M* = 51.58, *SD* = 10.74 | *t*(11) = 3.84, *p* = **.003**, *d* = 1.11 |
| | Visit 6 | *W*(12) = .969, *p* = .896 | *M* = 46.67, *SD* = 10.70 | |
| D-KEFS Verbal Fluency: Task 2 (Category Fluency) | Visit 1 | *W*(12) = .894, *p* = .131 | *M* = 46.50, *SD* = 7.49 | *t*(11) = 0, p = 1, *d* = 0 |
| | Visit 6 | *W*(12) = .939, *p* = .485 | *M* = 46.50, *SD* = 6.76 | |
| D-KEFS Verbal Fluency: Task 3 (Category Switching) | Visit 1 | *W*(12) = .826, *p* = **.019** | *M* = 13.83, *SD* = 3.84 | *t*(11) = .65, *p* = .529, *d* = .19 |
| | Visit 6 | *W*(12) = .924, *p* = .325 | *M* = 13.17, *SD* = 4.13 | |
| **Repeatable Battery for the Assessment of Neuropsychological Status (RBANS) Update: 12 individual tasks** | | | | |

| | | | | |
|---|---|---|---|---|
| List Learning | Visit 1 | *W*(12) = .944, *p* = .547 | *M* = 29.00, *SD* = 4.79 | *t*(11) = −.611, *p* = .554, *d* = −.18 |
| | Visit 6 | *W*(12) = .981, *p* = .987 | *M* = 30.00, *SD* = 5.34 | |
| Story Memory | Visit 1 | *W*(12) = .937, *p* = .462 | *M* = 17.83, *SD* = 2.95 | *t*(11) = −1.184, *p* = .261, *d* = −.34 |
| | Visit 6 | *W*(12) = .934, *p* = .429 | *M* = 19.25, *SD* = 2.63 | |
| Figure Copy | Visit 1 | *W*(12) = .930, *p* = .381 | *M* = 17.75, *SD* = 1.55 | *t*(11) = .897, *p* = .389, *d* = .26 |
| | Visit 6 | *W*(12) = .942, *p* = .518 | *M* = 17.25, *SD* = 2.01 | |
| Line Orientation | Visit 1 | *W*(12) = .838, *p* = **.026** | *Mdn* = 18.50, *IQR* = 4.00 | *Z* = .284, *p* = .776, *r* = .08 |
| | Visit 6 | *W*(12) = .817, *p* = **.015** | *Mdn* = 18.50, *IQR* = 3.00 | |
| Picture Naming | Visit 1 | *W*(12) = .465, *p* < **.001** | *Mdn* = 10.00, *IQR* = 0.00 | *Z* = 0, *p* = 1, *r* = 0 |
| | Visit 6 | *W*(12) = .327, *p* < **.001** | *Mdn* = 10.00, *IQR* = 0.00 | |
| Semantic Fluency | Visit 1 | *W*(12) = .909, *p* = .210 | *M* = 24.50, *SD* = 5.67 | *t*(11) = .930, *p* = .372, *d* = .27 |
| | Visit 6 | *W*(12) = .896, *p* = .139 | *M* = 22.58, *SD* = 4.89 | |
| Digit Span | Visit 1 | *W*(12) = .910, *p* = .211 | *M* = 12.25, *SD* = 3.11 | *t*(11) = −.312, *p* = .761, *d* = −.09 |
| | Visit 6 | *W*(12) = .892, *p* = .127 | *M* = 12.42, *SD* = 2.47 | |
| Coding | Visit 1 | *W*(11) = .945, *p* = .585 | *M* = 52.00, *SD* = 7.23 | *t*(11) = .309, *p* = .764, *d* = .68 |
| | Visit 6 | *W*(11) = .960, *p* = .768 | *M* = 51.45, *SD* = 8.92 | |
| List Recall | Visit 1 | *W*(12) = .919, *p* = .280 | *M* = 6.92, *SD* = 2.31 | t(11) = 1.316, *p* = .215, *d* = .38 |
| | Visit 6 | *W*(12) = .925, *p* = .329 | *M* = 5.75, *SD* = 3.31 | |
| List Recognition | Visit 1 | *W*(12) = .779, *p* = **.005** | *Mdn* = 19.50, *IQR* = 2.00 | *Z* = 1.354, *p* = .176, *r* = .39 |
| | Visit 6 | *W*(12) = .814, *p* = **.014** | *Mdn* = 18.00, *IQR* = 3.00 | |
| Story Recall | Visit 1 | *W*(12) = .905, *p* = .185 | *M* = 9.67, *SD* = 1.97 | *t*(11) = −.613, *p* = .552, *d* = −.18 |
| | Visit 6 | *W*(12) = .888, *p* = .113 | *M* = 10.08, *SD* = 1.78 | |
| Figure Recall | Visit 1 | *W*(12) = .956, *p* = .722 | *M* = 14.67, *SD* = 3.63 | *t*(11) = −.823, *p* = .428, *d* = −.24 |
| | Visit 6 | *W*(12) = .964, *p* = .845 | *M* = 15.33, *SD* = 2.74 | |
| **Repeatable Battery for the Assessment of Neuropsychological Status (RBANS) Update: 5 cognitive domains** | | | | |
| Immediate Memory | Visit 1 | *W*(12) = .960, *p* = .788 | *M* = 98.83, *SD* = 16.56 | *t*(11) = −.531, *p* = .606, *d* = −.15 |
| | Visit 6 | *W*(12) = .935, *p* = .440 | *M* = 102.33, *SD* = 12.43 | |
| Visuospatial/Constructional Abilities | Visit 1 | *W*(12) = .937, *p* = .459 | *M* = 97.17, *SD* = 12.83 | *t*(11) = .835, *p* = .421, *d* = .24 |
| | Visit 6 | *W*(12) = .929, *p* = .370 | *M* = 93.50, *SD* = 16.09 | |
| Language | Visit 1 | *W*(12) = .913, *p* = .234 | *M* = 106.17, *SD* = 12.66 | |

| | | | | |
|---|---|---|---|---|
| | Visit 6 | *W*(12) = .907, *p* = .193 | *M* = 101.25, *SD* = 12.02 | *t*(11) = 1.121, *p* = .286, *d* = .32 |
| Attention | Visit 1 | *W*(11) = .932, *p* = .436 | *M* = 104.82, *SD* = 18.45 | *t*(10) = −.278, *p* = .787, *d* = .51 |
| | Visit 6 | *W*(11) = .954, *p* = .700 | *M* = 105.64, *SD* = 14.33 | |
| Delayed Memory | Visit 1 | *W*(12) = .956, *p* = .729 | *M* = 94.75, *SD* = 20.05 | *t*(11) = .758, *p* = .464, *d* = .22 |
| | Visit 6 | *W*(12) = .947, *p* = .591 | *M* = 90.58, *SD* = 19.70 | |
| **Repeatable Battery for the Assessment of Neuropsychological Status (RBANS) Update: Total scores** | | | | |
| Sum of Index Score | Visit 1 | *W*(11) = .950, *p* = .649 | *M* = 503.82, *SD* = 51.36 | *t*(10) = .834, *p* = .424, *d* = .85 |
| | Visit 6 | *W*(11) = .945, *p* = .578 | *M* = 491.45, *SD* = 39.56 | |
| Total Scaled Score | Visit 1 | *W*(11) = .934, *p* = .450 | *M* = 100.82, *SD* = 15.15 | *t*(10) = .860, *p* = .410, *d* = .86 |
| | Visit 6 | *W*(11) = .961, *p* = .784 | *M* = 97.18, *SD* = 10.16 | |
| **Trail Making Test (TMT)** | | | | |
| TMT: Part A | Visit 1 | *W*(11) = .901, *p* = .192 | *Mdn* = 21, *IQR* = 15.00 | *Z* = 1.168, *p* = .243, *r* = .35 |
| | Visit 6 | *W*(11) = .658, *p* < **.001** | *Mdn* = 18, *IQR* = 10.00 | |
| TMT: Part B | Visit 1 | *W*(11) = .949, *p* = .635 | *M* = 56.55, *SD* = 11.27 | *t*(10) = 1.53, *p* = .157, *d* = .43 |
| | Visit 6 | *W*(11) = .984, *p* = .985 | *M* = 47.09, *SD* = 16.23 | |
| **Effort for Expenditure for Rewards Task (EEfRT)** | | | | |
| EEfRT | Visit 1 | *W*(9) = .129, *p* = .704 | *M* = .35, *SD* = .16 | *t*(8) = −1.12, *p* = .295, *d* = −.37 |
| | Visit 6 | *W*(9) = .145, *p* = .621 | *M* = .42, *SD* = .17 | |
| **Beck's Depression Inventory–II (BDI-II)** | | | | |
| BDI-II | Visit 1 | *W*(12) = .780, *p* = .006 | *Mdn* = 5.50, *IQR* = 7.00 | *Z* = 1.901, *p* = .057, *r* = .55 |
| | Visit 6 | *W*(12) = .774, *p* = .005 | *Mdn* = 3.00, *IQR* = 7.00 | |
| **Visual Analog Mood Scale (VAM-S)** | | | | |
| VAM-S | Visit 2 | *W*(12) = .937, *p* = .466 | *M* = 7.23, *SD* = .99 | Mauchly's test: $\chi^2(5) = 14.70$, *p* = **.012**, Greenhouse-Geisser correction: ε = .518, RM ANOVA: *F*(1.56, 17.10) = 1.40, *p* = .268, $\eta_p^2$ = .11 |
| | Visit 3 | *W*(12) = .947, *p* = .600 | *M* = 6.98, *SD* = 1.97 | |
| | Visit 4 | *W*(12) = .970, *p* = .906 | *M* = 7.72, *SD* = 1.24 | |
| | Visit 5 | *W*(12) = .969, *p* = .898 | *M* = 7.14, *SD* = 1.57 | |

**Table S4.** The output of post hoc tests (paired-samples *t*-tests or Wilcoxon signed-rank tests), uncorrected and following the correction for multiple comparisons using the false discovery rate (FDR) method (Benjamini & Hochberg, 1995).

| | **Compared visits** | **Uncorrected *p*-value** | **FDR-corrected *p*-value** |
|---|---|---|---|
| | | **Vitals during AIH** | |
| max HR | Visit 2 *vs* Visit 3 | $t(11) = -1.271$, $p = .230$, $d = -.37$ | $p = .276$ |
| | Visit 2 *vs* Visit 4 | $t(11) = -1.784$, $p = .102$, $d = -.52$ | $p = .192$ |
| | Visit 2 *vs* Visit 5 | $t(11) = -3.618$, $p =$ **.004**, $d = -1.04$ | $p =$ **.012** |
| | Visit 3 *vs* Visit 4 | $t(11) = -.217$, $p = .832$, $d = -.06$ | $p = .832$ |
| | Visit 3 *vs* Visit 5 | $t(11) = -1.644$, $p = .128$, $d = -.48$ | $p = .192$ |
| | Visit 4 *vs* Visit 5 | $t(11) = -3.943$, $p =$ **.002**, $d = -1.14$ | $p =$ **.012** |
| min $SpO_2$ | Visit 2 *vs* Visit 3 | $t(11) = 2.570$, $p =$ **.026**, $d = .74$ | $p =$ **.026** |
| | Visit 2 *vs* Visit 4 | $t(11) = 6.134$, $p <$ **.001**, $d = 1.77$ | $p =$ **.001** |
| | Visit 2 *vs* Visit 5 | $t(11) = 13.075$, $p <$ **.001**, $d = 3.77$ | $p =$ **.001** |
| | Visit 3 *vs* Visit 4 | $t(11) = 5.631$, $p <$ **.001**, $d = 1.63$ | $p =$ **.001** |
| | Visit 3 *vs* Visit 5 | $t(11) = 16.659$, $p <$ **.001**, $d = 4.81$ | $p =$ **.001** |
| | Visit 4 *vs* Visit 5 | $t(11) = 12.775$, $p <$ **.001**, $d = 3.69$ | $p =$ **.001** |
| | | **Finger Tapping Test** | |
| Finger Tapping Test, right hand | Visit 1 *vs* Visit 2 | $Z = -1.824$, $p = .068$, $r = -.55$ | $p = .113$ |
| | Visit 1 *vs* Visit 3 | $Z = -2.191$, $p =$ **.028**, $r = -.66$ | $p = .053$ |
| | Visit 1 *vs* Visit 4 | $Z = -2.936$, $p =$ **.003**, $r = -.89$ | $p =$ **.019** |
| | Visit 1 *vs* Visit 5 | $Z = -2.934$, $p =$ **.003**, $r = -.88$ | $p =$ **.019** |
| | Visit 1 *vs* Visit 6 | $Z = -2.845$, $p =$ **.004**, $r = -.86$ | $p =$ **.019** |
| | Visit 2 *vs* Visit 3 | $Z = -.711$, $p = .477$, $r = -.21$ | $p = .511$ |
| | Visit 2 *vs* Visit 4 | $Z = -2.192$, $p =$ **.028**, $r = -.66$ | $p = .053$ |
| | Visit 2 *vs* Visit 5 | $Z = -2.293$, $p =$ **.022**, $r = -.69$ | $p = .053$ |
| | Visit 2 *vs* Visit 6 | $Z = -1.646$, $p = .100$, $r = -.50$ | $p = .150$ |
| | Visit 3 *vs* Visit 4 | $Z = -1.274$, $p = .203$, $r = -.38$ | $p = .276$ |
| | Visit 3 *vs* Visit 5 | $Z = -2.805$, $p =$ **.005**, $r = -.85$ | $p =$ **.019** |
| | Visit 3 *vs* Visit 6 | $Z = -2.627$, $p =$ **.009**, $r = -.79$ | $p =$ **.027** |

| | | | |
|---|---|---|---|
| | Visit 4 *vs* Visit 5 | *Z* = −1.224, *p* = .221, *r* = −.37 | *p* = .276 |
| | Visit 4 *vs* Visit 6 | *Z* = −1.023, *p* = .306, *r* = −.31 | *p* = .353 |
| | Visit 5 *vs* Visit 6 | *Z* = −.306, *p* = .759, *r* = −.09 | *p* = .759 |
| Finger Tapping Test, left hand | Visit 1 *vs* Visit 2 | *t*(10) = −.880, *p* = .400, *d* = −.27 | *p* = .500 |
| | Visit 1 *vs* Visit 3 | *t*(10) = −1.346, *p* = .208, *d* = −.41 | *p* = .314 |
| | Visit 1 *vs* Visit 4 | *t*(10) = −1.343, *p* = .209, *d* = −.41 | *p* = .314 |
| | Visit 1 *vs* Visit 5 | *t*(10) = −3.415, *p* = **.007**, *d* = −1.03 | *p* = .053 |
| | Visit 1 *vs* Visit 6 | *t*(10) = −3.538, *p* = **.005**, *d* = −1.07 | *p* = .053 |
| | Visit 2 *vs* Visit 3 | *t*(10) = −.746, *p* = .473, *d* = −.23 | *p* = .546 |
| | Visit 2 *vs* Visit 4 | *t*(10) = −1.056, *p* = .316, *d* = −.32 | *p* = .431 |
| | Visit 2 *vs* Visit 5 | *t*(10) = −3.084, *p* = **.012**, *d* = −.93 | *p* = .060 |
| | Visit 2 *vs* Visit 6 | *t*(10) = −2.238, *p* = **.049**, *d* = −.68 | *p* = .105 |
| | Visit 3 *vs* Visit 4 | *t*(10) = −.365, *p* = .723, *d* = −.11 | *p* = .741 |
| | Visit 3 *vs* Visit 5 | *t*(10) = −2.540, *p* = **.029**, *d* = −.77 | *p* = .103 |
| | Visit 3 *vs* Visit 6 | *t*(10) = −2.340, *p* = **.041**, *d* = −.71 | *p* = .103 |
| | Visit 4 *vs* Visit 5 | *t*(10) = −2.381, *p* = **.039**, *d* = −.72 | *p* = .103 |
| | Visit 4 *vs* Visit 6 | *t*(10) = −1.769, *p* = .107, *d* = −.53 | *p* = .201 |
| | Visit 5 *vs* Visit 6 | *t*(10) = −.340, *p* = .741, *d* = −.11 | *p* = .741 |
| **Grooved Pegboard Test** | | | |
| Grooved Pegboard Test, right hand | Visit 1 *vs* Visit 2 | *Z* = 2.652, *p* = **.008**, *r* = .84 | *p* = **.034** |
| | Visit 1 *vs* Visit 3 | *Z* = 2.383, *p* = **.017**, *r* = .75 | *p* = .051 |
| | Visit 1 *vs* Visit 4 | *Z* = 2.603, *p* = **.009**, *r* = .82 | *p* = **.034** |
| | Visit 1 *vs* Visit 5 | *Z* = 2.807, *p* = **.005**, *r* = .89 | *p* = **.034** |
| | Visit 1 *vs* Visit 6 | *Z* = 2.601, *p* = **.009**, *r* = .82 | *p* = **.034** |
| | Visit 2 *vs* Visit 3 | *Z* = .297, *p* = .767, *r* = .09 | *p* = .885 |
| | Visit 2 *vs* Visit 4 | *Z* = .408, *p* = .683, *r* = .13 | *p* = .854 |
| | Visit 2 *vs* Visit 5 | *Z* = 1.721, *p* = .085, *r* = .54 | *p* = .182 |
| | Visit 2 *vs* Visit 6 | *Z* = 1.147, *p* = .251, *r* = .36 | *p* = .377 |
| | Visit 3 *vs* Visit 4 | *Z* = .103, *p* = .918, *r* = .03 | *p* = .918 |
| | Visit 3 *vs* Visit 5 | *Z* = 1.614, *p* = .106, *r* = .51 | *p* = .199 |

| | | | |
|---|---|---|---|
| | Visit 3 *vs* Visit 6 | *Z* = .919, *p* = .358, *r* = .29 | *p* = .488 |
| | Visit 4 *vs* Visit 5 | *Z* = 1.889, *p* = .059, *r* = .60 | *p* = .148 |
| | Visit 4 *vs* Visit 6 | *Z* = 1.187, *p* = .235, *r* = .38 | *p* = .377 |
| | Visit 5 *vs* Visit 6 | *Z* = .140, *p* = .889, *r* = .04 | *p* = .918 |
| Grooved Pegboard Test, left hand | Visit 1 *vs* Visit 2 | *t*(9) = 3.466, p = **.007**, *d* = 1.10 | *p* = **.026** |
| | Visit 1 *vs* Visit 3 | *t*(9) = 2.790, *p* = **.021**, *d* = .88 | *p* = .063 |
| | Visit 1 *vs* Visit 4 | *t*(9) = 4.143, *p* = **.003**, *d* = 1.31 | *p* = **.015** |
| | Visit 1 *vs* Visit 5 | *t*(9) = 3.943, *p* = **.003**, *d* = 1.25 | *p* = **.015** |
| | Visit 1 *vs* Visit 6 | *t*(9) = 4.143, *p* = **.003**, *d* = 1.31 | *p* = **.015** |
| | Visit 2 *vs* Visit 3 | *t*(9) = −.098, *p* = .924, *d* = −.03 | *p* = .924 |
| | Visit 2 *vs* Visit 4 | *t*(9) = 1.903, *p* = .089, *d* = .60 | *p* = .223 |
| | Visit 2 *vs* Visit 5 | *t*(9) = 1.438, *p* = .184, *d* = .46 | *p* = .305 |
| | Visit 2 *vs* Visit 6 | *t*(9) = 1.427, *p* = .187, *d* = .45 | *p* = .305 |
| | Visit 3 *vs* Visit 4 | *t*(9) = 1.606, *p* = .143, *d* = .51 | *p* = .305 |
| | Visit 3 *vs* Visit 5 | *t*(9) = 1.374, *p* = .203, *d* = .44 | *p* = .305 |
| | Visit 3 *vs* Visit 6 | *t*(9) = 1.223, *p* = .252, *d* = .39 | *p* = .344 |
| | Visit 4 *vs* Visit 5 | *t*(9) = −.190, *p* = .853, *d* = −.06 | *p* = .914 |
| | Visit 4 *vs* Visit 6 | *t*(9) = −.736, *p* = .480, *d* = −.23 | *p* = .600 |
| | Visit 5 *vs* Visit 6 | *t*(9) = −.264, *p* = .798, *d* = −.08 | *p* = .914 |

### Supplementary Figures

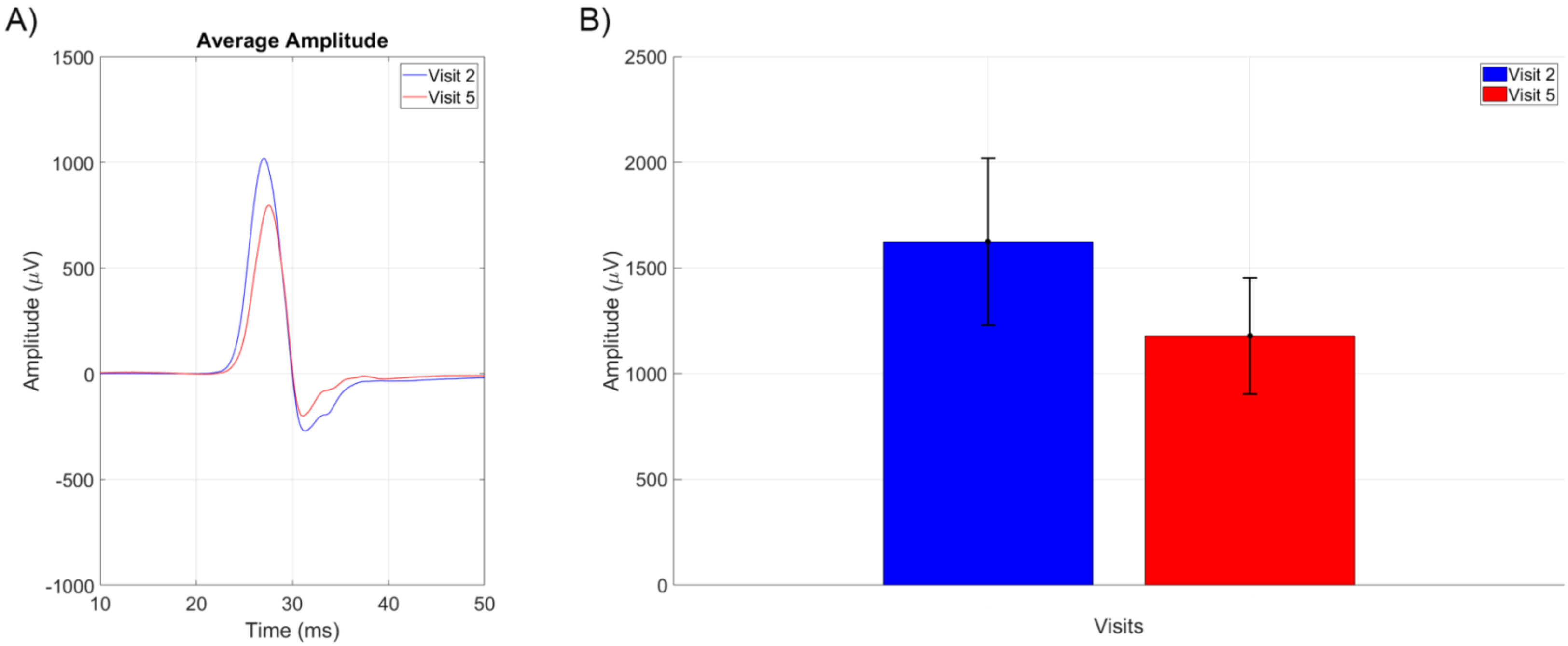

**Figure S1.** Group-level corticospinal responses to transcranial magnetic stimulation (TMS) of the motor hand area recorded from the right first dorsal interosseous (FDI) muscle. **(A)** Average motor evoked potential (MEP) time courses across nine participants. **(B)** Group-level peak-to-peak MEP amplitudes; error bars represent the standard error of the mean (SEM). The MEPs decreased in Visit 5; however, the changes were not significant.

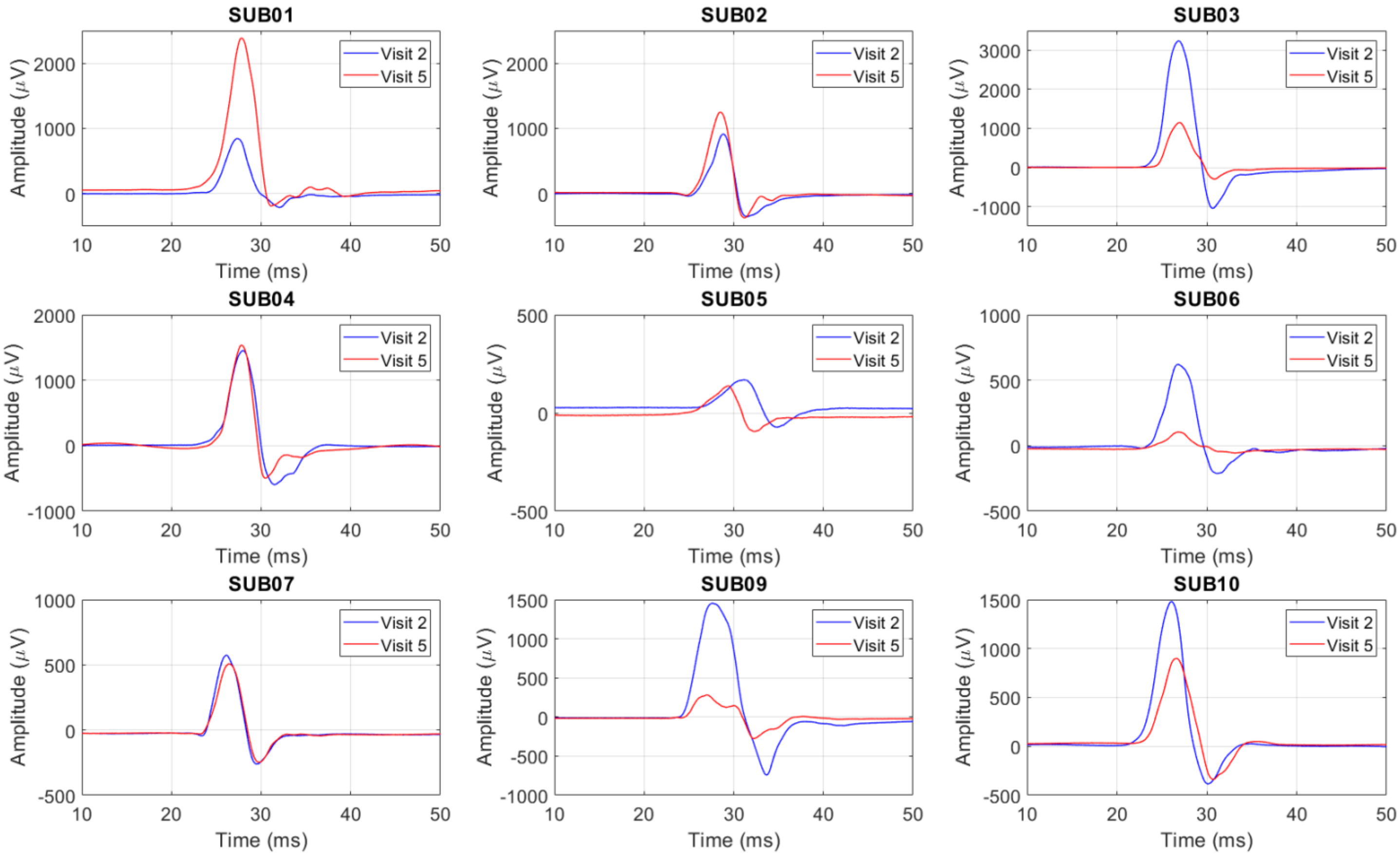

**Figure S2.** Subject-level corticospinal responses to transcranial magnetic stimulation (TMS) of the motor hand area recorded from the right first dorsal interosseous (FDI) muscle. The direction of changes in motor evoked potential (MEP) time courses varied across participants.
